# Evaluation of improvement training for cancer MDT members

**DOI:** 10.64898/2026.11.14.26367953

**Authors:** Ayah Atiyah, Benjamin W Lamb, Tayana Soukup

## Abstract

**Background:** Multidisciplinary team (MDT) meetings are central to cancer care. National policy, including the 2026 National Cancer Plan for England, expects MDTs to improve how they work. Although validated tools assess MDT performance, little is known about training in their use. We evaluated structured MDT improvement training delivered across two UK Cancer Alliances.

**Methods:** Three one-day courses, delivered in 2019, combined teaching and simulation using MDT assessment tools, including the Metric for the Observation of Decision-making (MDT-MODe). Participants completed a 14-item Likert-scale survey (1 = strongly disagree to 5 = strongly agree) and open-ended questions. Responses were summarised descriptively; subgroup comparisons used chi-squared tests with Bonferroni correction. Free-text responses were analysed thematically by two coders.

**Results:** Forty-eight MDT members responded (26 doctors, 10 nurses, 10 administrative staff). Twelve of 14 items scored a median of 4 (agree); understanding the information scored a 4.5. Support for mandatory training was more modest (median 3.5). After Bonferroni correction, significant differences by grade remained for knowledge of the MDT-metric (adjusted p = 0.001) and MDT-MODe instrument (adjusted p < 0.001). Strengths included teaching (n = 14), utility of MDT-MODe (n = 10) and networking (n = 9). Weaknesses included poor audio-visual quality (n = 18) and difficulty identifying speakers (n = 8).

**Conclusions:** Expert-led, simulation-based MDT improvement training was well received across professional groups. Although delivered in 2019, it addresses skills central to national MDT reform. Future courses should improve audio-visual quality and offer implementation support. Pre–post evaluation and longer-term follow-up are needed.

## 1 Background

Multidisciplinary teams (MDTs) represent the gold standard for the delivery of cancer care, both within the United Kingdom and internationally.[1] Central to this process are the multidisciplinary team meetings (MDMs), where treatment recommendations are formulated collaboratively. MDMs typically bring together histopathologists, radiologists, surgeons, cancer nurse specialists, research nurses, radiation oncologists, and medical oncologists, and are convened on a weekly basis.[2] However, the implementation and scope of MDMs can vary considerably, both between countries and within regions, ranging from the discussion of only complex cases to the systematic review of all newly diagnosed patients.[3]

Cancer Alliances in the UK are regional bodies that support cancer multidisciplinary teams (MDTs) to implement national health service policy by coordinating collaboration, overseeing performance and workforce needs among local providers.[2] Cancer alliances are required to facilitate improvements in MDT working with the aim of ensuring consistent, effective, and equitable MDT functioning across local systems. This has been achieved through a variety of quality improvement and innovation initiatives.[4–6]

Over the past decade, a growing body of improvement research has focused on enhancing the quality and effectiveness of cancer MDTs. This work has culminated in the development of a suite of tools designed to support clinicians, administrators, and researchers in objectively assessing team performance.[7] These tools are increasingly employed within audits and quality improvement initiatives to streamline meetings and ultimately enhance patient care delivery.[8]

To date, however, little is known about the impact of training in MDT improvement on the healthcare professionals to which it is delivered.

To facilitate the implementation of these improvements, we delivered a series of targeted training sessions at two UK cancer alliances.

The training content and structure

Three one-day courses were designed and conducted by experts in MDT improvement (BWL, TS), drawing upon backgrounds in teamworking, human factors, implementation science and clinical practice. Each session was attended by approximately 20 participants, comprising a diverse range of healthcare professionals including surgeons, oncologists, physicians, histopathologists, radiologists, specialist nurses, MDT coordinators, cancer managers, and others involved in MDT operations.

The training sessions were structured around small group teaching complemented by hands-on simulations using MDT assessment tools, fostering both theoretical understanding and practical application. The training combined didactic lectures, practical simulation exercises, and the provision of structured written materials. The session covered topics including: [could we add a few topics from the manual as examples of what we covered]

### 1.1 Aims and Objectives

The aim of this study was to evaluate the impact of MDT-focused training on participants’ knowledge, skills, and attitudes towards MDT functioning. Specifically, the study sought to:

- Assess participants’ perceptions of the quality and utility of the training provided
- Identify areas for potential improvement in future training sessions.

## 2 Methods

MDT training sessions were designed and conducted at multiple cancer centres across the United Kingdom by TS and BWL, targeting healthcare professionals integral to MDT operations, including surgeons, radiologists, physicians, cancer nurse specialists (CNSs), and administrative staff.

The training curriculum combined formal lectures with interactive components, such as video and audio-based case simulations, aimed at illustrating key principles of MDT assessment and decision-making. Participants engaged in both theoretical discussions and practical application of the MDT improvement tools during the sessions.

Following the completion of training, participants were invited to complete structured surveys. These surveys included a combination of Likert scale items and open-ended questions designed to capture both quantitative measures of satisfaction and qualitative insights into the perceived impact of the training on MDT understanding and practice.

Survey responses were manually transcribed into a secure database. Quantitative data were analysed using descriptive statistical methods to identify emerging trends. Qualitative responses were subjected to thematic analysis to extract recurring patterns and insights.

### 2.1 Study participants

A total number of 48 MDT members responded to the survey across 3 different training dates. This included doctors (consultants from different medical and surgical specialities), nurses and non-clinical staff (MDT coordinators).

### 2.2 Study design

Our evaluation consisted of two complementary components. The first component was quantitative, comprising a structured survey containing Likert-scale questions designed to assess participants’ satisfaction with the training content, delivery, and perceived impact on their understanding of MDT processes. The questions can be seen in Table 1 below.

**Table 1:** demographic characteristics of participants.

| Training date | Cohort size | % |
|---|---|---|
| 12/06/2019 | 21 | 43.8 |
| 19/06/2019 | 12 | 25.0 |
| 26/06/2019 | 15 | 31.3 |
| **Total** | **48** | **100.0** |
| Professional role | Number | % |
| Missing | 2 | 4.2 |
| Administrative | 10 | 20.8 |
| Nurse | 10 | 20.8 |
| Oncologist | 4 | 8.3 |
| Physician | 7 | 14.6 |
| Radiologist | 4 | 8.3 |
| Surgeon | 11 | 22.9 |
| Grade | Number | % |
| Missing | 3 | 6.3 |
| Band 4 | 3 | 6.3 |
| Band 5 | 4 | 8.3 |
| Band 6 | 2 | 4.2 |
| Band 7 | 8 | 16.7 |
| Band 8a | 2 | 4.2 |
| Consultant | 23 | 47.9 |
| Junior doctor | 3 | 6.3 |
| Speciality | Number | % |
| Missing | 2 | 4.2 |
| Breast | 7 | 14.6 |
| Cancer services | 5 | 10.4 |
| Colorectal | 5 | 10.4 |
| ENT | 2 | 4.2 |
| Gastroenterology | 1 | 2.1 |
| Gynaecology | 3 | 6.3 |
| Haematology | 2 | 4.2 |
| Head and neck | 3 | 6.3 |
| Lung | 6 | 12.5 |
| Medical oncology | 1 | 2.1 |
| Medicine | 1 | 2.1 |
| Oncology | 2 | 4.2 |
| Radiology | 3 | 6.3 |
| Skin | 2 | 4.2 |
| Upper gastrointestinal | 2 | 4.2 |
| Urology | 1 | 2.1 |
| MDT Lead | Number | % |
| Specified yes | 4 | 8.3 |
| Not specified | 44 | 91.7 |

The second component was qualitative, eliciting free-text responses from participants regarding their perceptions of the MDT training day and the MDT improvement tool, including aspects they found beneficial and areas where improvements could be made.

Free-text responses captured insights into strengths and weaknesses of both the training sessions and the tool, allowing an exploration of the participant experience. Likert-scale questions ranged from 1 (strongly disagree) to 5 (strongly agree), providing a quantitative measure of participants’ views across domains including training relevance, clarity, practical application, and overall satisfaction.

### 2.3 Qualitative and quantitative analysis

#### Qualitative analysis

48 MDT members responded to the survey. The analytical approach followed established standards in qualitative research to ensure validity and transparency.[27] First, responses to the survey were collated into a single document to form a transcript. These transcripts were then analyzed independently by two blinded coders (AA and BWL). The coders aimed to extract from the transcripts ‘themes’ in relation to each one of the three open-ended questions and to allocate all of participants’ responses to these themes.

Themes were tabulated, and verbatim quotes from respondents to validate and illustrate the themes were extracted. To simplify presentation themes reported by a very small minority of respondents (fewer than 5% of the total) are not presented in this paper, but are available from the corresponding author.

#### Quantitative analysis

SPSS IBM SPSS Statistics 24 was used to explore quantitative survey responses. Descriptive statistics were generated for each question. To assess differences in responses between professional groups, grades, tumour type and training sessions the Chi-Squared test was used. Each of the four demographic characteristics (training date, professional role, grade and tumour type) was compared across the 14 Likert-scale questions, giving 56 tests in total. Respondents with missing demographic data were excluded from the relevant comparison. To account for multiple testing, a Bonferroni correction was applied within each demographic characteristic, with the 14 comparisons for that characteristic treated as one family. A difference was considered statistically significant if the unadjusted p-value was below 0.0036 (0.05/14), equivalent to a Bonferroni-adjusted p-value (unadjusted p-value × 14, capped at 1) below 0.05. As a sensitivity analysis, all 56 comparisons were also treated as a single family (significance threshold 0.05/56 = 0.00089). Because the sample was small and several demographic characteristics included many categories, many contingency-table cells had expected counts below five. These subgroup comparisons were therefore regarded as exploratory and hypothesis-generating, and were interpreted cautiously.

## 3 Results

### 3.1 Participant characteristics

The participants were MDT members (N = 48) who attended the training across three course dates (details seen in Table 1 below). The cohort represented a range of professional groups, including administrative staff (n = 10), nurses (n = 10; ranging from Band 4 to Band 8a), and doctors (n = 26), comprising both junior doctors and consultants. Among the doctors, there were surgeons (n = 11), radiologists (n = 4), oncologists (n = 4), and physicians (n = 7). Two participants did not specify their professional role.

Participants reported working across a range of cancer specialties, covering breast (n = 7), upper gastrointestinal (n = 2), colorectal (n = 5), ENT and head and neck (n = 5), gynaecology (n = 3), lung (n = 6), skin (n = 2), and urology (n = 1) services.

### 3.2 Quantitative questions

Table 2 and Figures 1 and 2 display data regarding responses to quantitative survey questions.

**Figure 1:**
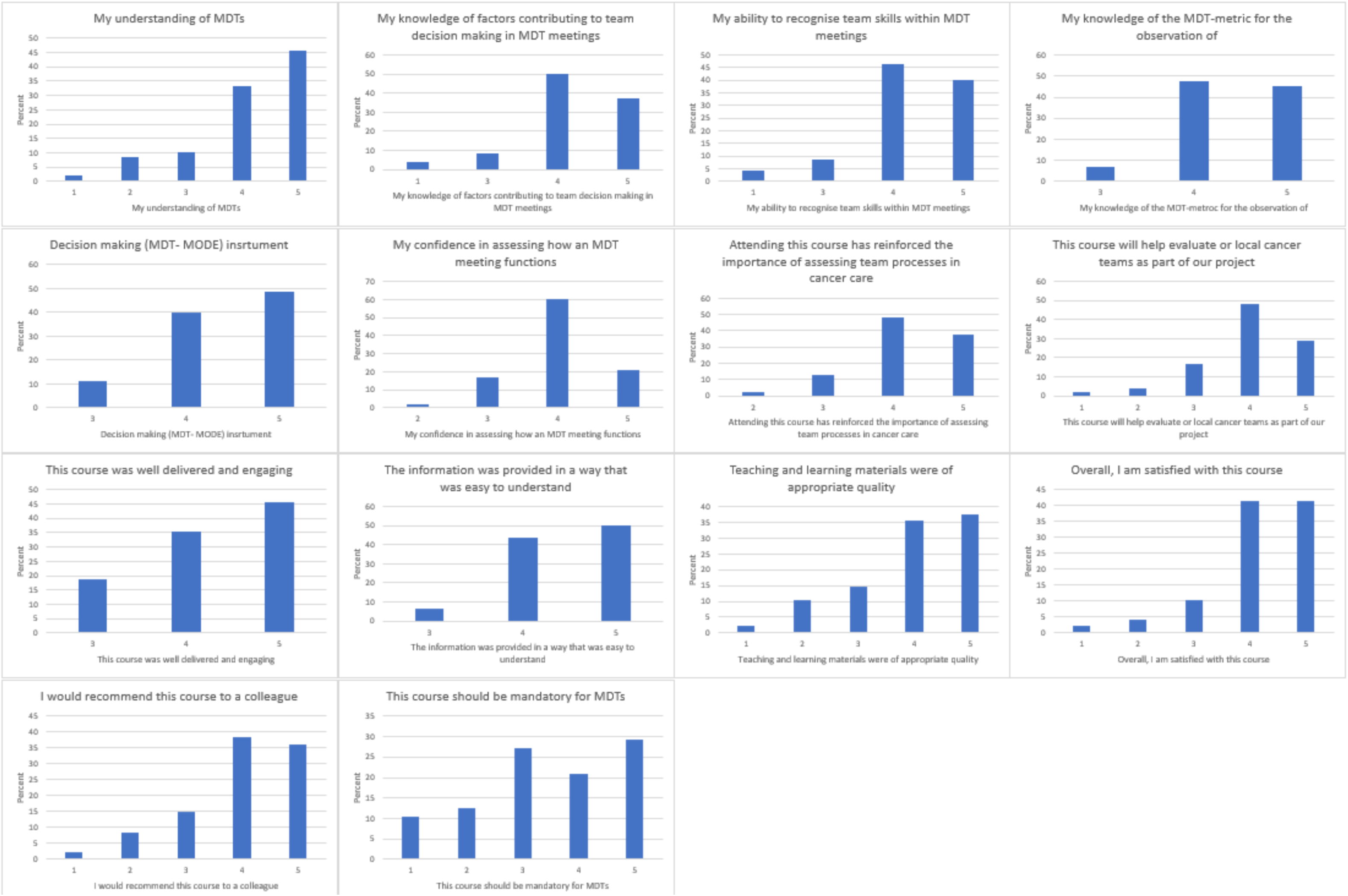
Bar graphs displaying data regarding level of agreement with statements about training.

**Figure 2:**
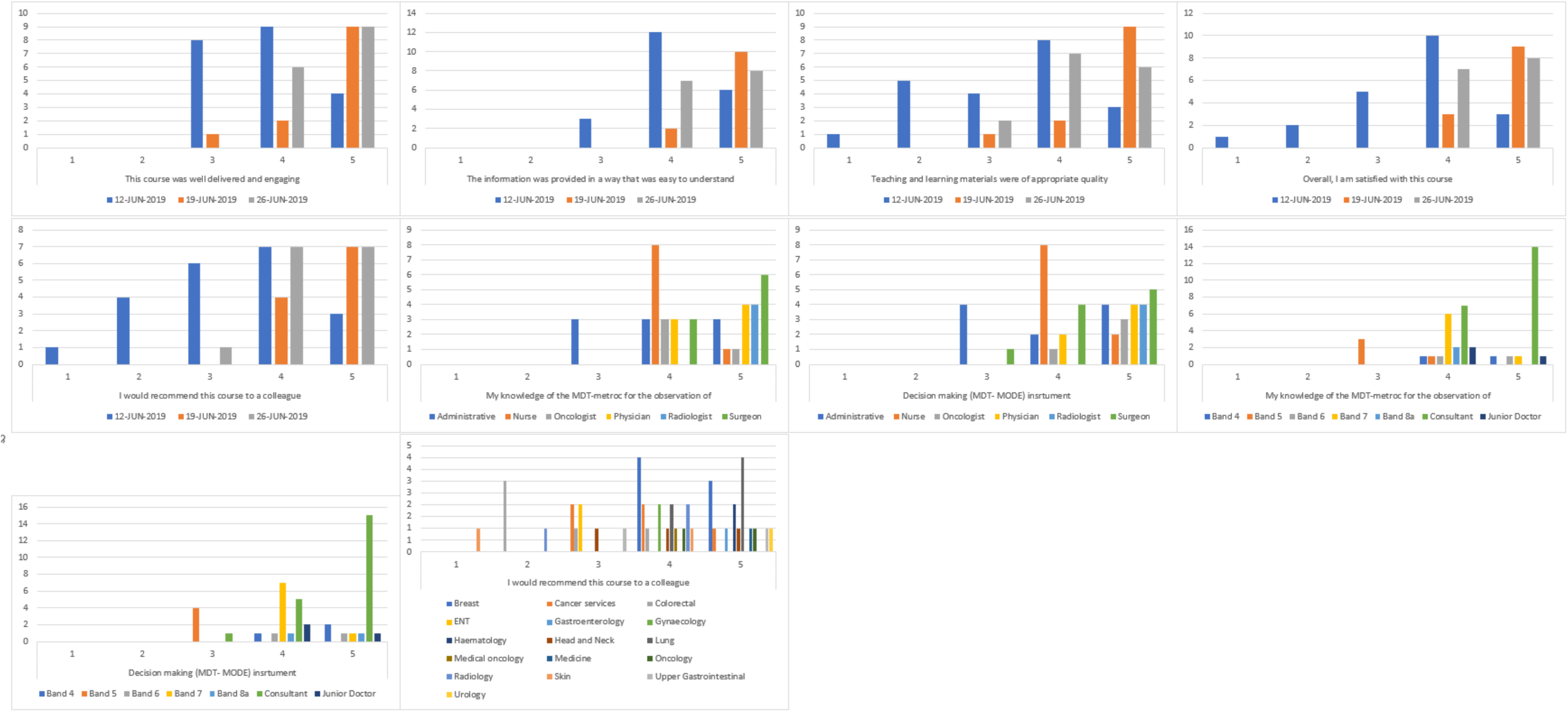
Bar graphs displaying differences between demographic groups regarding level of agreement with statements about training. The panels show the 10 comparisons with an unadjusted p-value below 0.05. Only the two comparisons according to grade (My knowledge of the MDT-metric; and for the observation of Decision making (MDT-MODE) instrument) remained statistically significant after Bonferroni correction.

**Table 2:** Quantitative evaluation questions and responses.

|  | descriptive |  |  |  | % responses |  |  |  |  | Date of Training |  |  |  | Role of respondent |  |  |  | Grade of respondent |  |  |  | Respondents’ MDT Tumour type |  |  |  |
|---|---|---|---|---|---|---|---|---|---|---|---|---|---|---|---|---|---|---|---|---|---|---|---|---|---|
|  | N | Med | Min | Max | 1 | 2 | 3 | 4 | 5 | Value | df | ***p*** | ***p***<sub>**adj**</sub> | Value | df | ***p*** | ***p***<sub>**adj**</sub> | Value | df | ***p*** | ***p***<sub>**adj**</sub> | Value | df | ***p*** | ***p***<sub>**adj**</sub> |
| My understanding of MDTs | 48 | 4 | 1 | 5 | 2.1 | 8.3 | 10.4 | 33.3 | 45.8 | 14.04 | 8 | 0.081 | 1.000 | 27.88 | 20 | 0.112 | 1.000 | 17.21 | 24 | 0.840 | 1.000 | 62.85 | 60 | 0.376 | 1.000 |
| My knowledge of factors contributing to team decision making in MDT meetings | 48 | 4 | 1 | 5 | 4.2 | 0 | 8.3 | 50 | 37.5 | 5.41 | 6 | 0.493 | 1.000 | 13.27 | 15 | 0.582 | 1.000 | 6.74 | 18 | 0.992 | 1.000 | 44.55 | 45 | 0.491 | 1.000 |
| My ability to recognise team skills within MDT meetings | 45 | 4 | 1 | 5 | 4.4 | 0 | 8.9 | 46.7 | 40 | 5.64 | 6 | 0.464 | 1.000 | 19.96 | 15 | 0.174 | 1.000 | 15.34 | 18 | 0.639 | 1.000 | 53.72 | 45 | 0.175 | 1.000 |
| My knowledge of the MDT-metric | 44 | 4 | 3 | 5 | 0 | 0 | 6.8 | 47.7 | 45.5 | 2.53 | 4 | 0.639 | 1.000 | 24.30 | 10 | 0.007 | 0.098 | 39.09 | 12 | <0.001 | **0.001\*** | 32.06 | 30 | 0.365 | 1.000 |
| for the observation of Decision making (MDT- MODE) instrument | 45 | 4 | 3 | 5 | 0 | 0 | 11.1 | 40 | 48.9 | 1.89 | 4 | 0.756 | 1.000 | 21.91 | 10 | 0.016 | 0.224 | 44.71 | 12 | <0.001 | **<0.001\*** | 39.78 | 28 | 0.069 | 0.966 |
| My confidence in assessing how an MDT meeting functions | 48 | 4 | 2 | 5 | 0 | 2.1 | 16.7 | 60.4 | 20.8 | 6.60 | 6 | 0.360 | 1.000 | 17.45 | 15 | 0.293 | 1.000 | 18.91 | 18 | 0.397 | 1.000 | 55.19 | 45 | 0.142 | 1.000 |
| Attending this course has reinforced the importance of assessing team processes in cancer care | 48 | 4 | 2 | 5 | 0 | 2.1 | 12.5 | 47.9 | 37.5 | 8.36 | 6 | 0.213 | 1.000 | 8.16 | 15 | 0.917 | 1.000 | 25.05 | 18 | 0.124 | 1.000 | 34.35 | 45 | 0.876 | 1.000 |
| This course will help evaluate or local cancer teams as part of our project | 48 | 4 | 1 | 5 | 2.1 | 4.2 | 16.7 | 47.9 | 29.2 | 11.80 | 8 | 0.159 | 1.000 | 14.36 | 15 | 0.499 | 1.000 | 15.02 | 18 | 0.661 | 1.000 | 69.45 | 60 | 0.189 | 1.000 |
| This course was well delivered and engaging | 48 | 4 | 3 | 5 | 0 | 0 | 18.8 | 35.4 | 45.8 | 15.48 | 4 | 0.004 | 0.056 | 8.17 | 10 | 0.613 | 1.000 | 14.35 | 12 | 0.279 | 1.000 | 32.33 | 30 | 0.352 | 1.000 |
| The information was provided in a way that was easy to understand | 48 | 4.5 | 3 | 5 | 0 | 0 | 6.3 | 43.8 | 50 | 11.39 | 4 | 0.023 | 0.322 | 4.82 | 10 | 0.903 | 1.000 | 5.88 | 12 | 0.922 | 1.000 | 31.82 | 30 | 0.376 | 1.000 |
| Teaching and learning materials were of appropriate quality | 48 | 4 | 1 | 5 | 2.1 | 10.4 | 14.6 | 35.4 | 37.5 | 17.66 | 8 | 0.024 | 0.336 | 10.74 | 15 | 0.771 | 1.000 | 13.41 | 18 | 0.767 | 1.000 | 48.17 | 45 | 0.346 | 1.000 |
| Overall, I am satisfied with this course | 48 | 4 | 1 | 5 | 2.1 | 4.2 | 10.4 | 41.7 | 41.7 | 18.82 | 8 | 0.016 | 0.224 | 19.35 | 20 | 0.499 | 1.000 | 8.53 | 24 | 0.998 | 1.000 | 67.30 | 60 | 0.241 | 1.000 |
| I would recommend this course to a colleague | 47 | 4 | 1 | 5 | 2.1 | 8.5 | 14.9 | 38.3 | 36.2 | 17.10 | 8 | 0.029 | 0.406 | 23.84 | 20 | 0.250 | 1.000 | 10.97 | 24 | 0.989 | 1.000 | 81.10 | 60 | 0.036 | 0.504 |
| This course should be mandatory for MDTs | 48 | 3.5 | 1 | 5 | 10.4 | 12.5 | 27.1 | 20.8 | 29.2 | 13.84 | 8 | 0.086 | 1.000 | 10.38 | 20 | 0.961 | 1.000 | 22.33 | 24 | 0.559 | 1.000 | 64.82 | 60 | 0.312 | 1.000 |

Med, median; Min, minimum; Max, maximum; df, degrees of freedom. Responses: 1 = strongly disagree to 5 = strongly agree. Value, df and p: Pearson chi-squared tests comparing responses across training dates (3 groups), professional roles (6), grades (7) and tumour types (16); missing demographic data excluded. p, unadjusted p-value; p<sub>adj</sub>, Bonferroni-adjusted p-value (p × 14, capped at 1) for the 14 questions within each demographic characteristic. \*Significant after Bonferroni correction (p<sub>adj</sub> < 0.05; equivalent to unadjusted p < 0.0036).

Completion rate was high with 10 of 14 questions were answered by all participants; the lowest number of responses to any question was 44. 12 of 14 questions had a median response of 4/5, signifying ‘agree’. One question, “The information was provided in a way that was easy to understand” had a median score of 4.5, signifying ‘agree/strongly agree’. One question, “This course should be mandatory for MDTs” received a median score of 3.5, signifying ‘neither agree nor disagree/agree’.

Before correction for multiple testing, 10 of the 56 between-group comparisons had an unadjusted p-value below 0.05 (Table 2; Figure 2). Five related to the date of training, for the questions: This course was well delivered and engaging; The information was provided in a way that was easy to understand; Teaching and learning materials were of appropriate quality; Overall, I am satisfied with this course; and I would recommend this course to a colleague. Two related to professional role and two to grade, for the questions: My knowledge of the MDT-metric; and for the observation of Decision making (MDT-MODE) instrument. One related to tumour type, for the question: I would recommend this course to a colleague.

After Bonferroni correction, only the differences according to grade remained statistically significant: My knowledge of the MDT-metric (χ² = 39.09, df = 12, adjusted p = 0.001); and for the observation of Decision making (MDT-MODE) instrument (χ² = 44.71, df = 12, adjusted p < 0.001). Both differences also remained significant when all 56 comparisons were treated as a single family (adjusted p = 0.006 and p < 0.001, respectively). Figure 2 suggests that consultants most frequently strongly agreed with these statements, whereas Band 5 respondents more often gave neutral responses; however, given the small number of respondents in several grade categories, these findings should be interpreted with caution. None of the differences according to training date, professional role or tumour type remained statistically significant after correction (adjusted p ≥ 0.056; Table 2). Visual inspection of Figure 2 suggests that scores were lower for the first day of training, but this observation is descriptive and should not be interpreted as a statistically significant difference. Because of the small sample size, further pairwise comparisons between specific groups were not undertaken.

### 3.3 Thematic analysis of free text responses

Table 3: themes of free text responses

**Table 3:** Thematic analysis of qualitative responses regarding training.

| **Theme** | **N** | **Representative Quote** | **Role** | **Grade** | **Specialty** |
|---|---|---|---|---|---|
| **Strengths** |  |  |  |  |  |
| Quality of presentation/teaching | 14 | "Well presented, good visuals" | Surgeon | Consultant | Breast |
| MDT MODE tool utility | 10 | "Identifying current areas of strengths and weaknesses within our team. Affirming current good practices and identifying how we can improve" | Nurse | Band 8a | Head and Neck |
| Networking and collaboration | 9 | "Good opportunity to meet colleagues from the region and discuss ideas/ strengths for managing care load/ MDT/ cancer patients" | Surgeon | Consultant | Colorectal |
| Practical/hands-on learning | 7 | "Identifying current areas of strengths and weaknesses within our team. Affirming current good practices and identifying how we can improve..." | Nurse | Band 8a | Head and Neck |
| Understanding MDT processes | 6 | "Understanding what goes into an MDT, the need for good communication between all parties" | Admin. | Band 5 | Cancer services |
| Positive overall feedback | 6 | "Excellent speakers and a great insight into MDT workings" | Nurse | Band 8a | Head and Neck |
| Research evidence base | 5 | "The enormous body of research has provided much needed insight into the process of MDTs to help improve performance" | Physician | Consultant | Gastro. |
| Course materials/resources | 3 | "Great to have a practical element/ experience using MDT MODE. Having a manual to take away" | Nurse | Band 7 | Upper GI |
| **Weaknesses** |  |  |  |  |  |
| Poor video/audio quality | 18 | "Better quality videos" | Nurse | Band 8a | Head and Neck |
| Difficulty identifying speakers in videos | 8 | "Video- sound needs improving, maybe use subtitles + show who’s speaking" | Admin. | Band 5 | Cancer services |
| Lack of practical implementation guidance | 4 | "More of a focus on practical strategies for improvement - what to do with the info when you've collected it" | Oncologist | Junior Doctor | Oncology |
| Repetitive/monotonous content | 3 | "[assessments] Became a bit monotonous towards the end" | Nurse | Band 8a | Head and Neck |
| Session too long | 3 | "The session could have been shortened" | Surgeon | Consultant | Colorectal |
| Limited scope of content | 3 | "Team skills within meetings could be touched upon more. Apart from information and contribution by team members there were no other factors discussed at depth" | Radiologist | Consultant | Radiology |
| Tool limitations/concerns | 3 | "To provide fair assessment of specialty contributions, the assessor needs to be sufficiently familiar with the normal treatment pathways to be able to judge whether 'lack of participation' is actually entirely appropriate. MDT MODE does not allow for this..." | Surgeon | Consultant | Colorectal |
| **Additional Comments** |  |  |  |  |  |
| Desire for follow-up/implementation support | 4 | "Good course, very informative. Might be nice to slightly more frequently done to discuss any changes/ improvements in MDTs" | Admin. | Band 5 | Cancer services |
| Request for broader content coverage | 4 | "Good idea but perhaps more information/ assessment of communication/ barriers to communication, adequate time allocation and resources" | Surgeon | Consultant | Colorectal |

Free-text responses were thematically analysed to explore participants’ perceptions of the MDT training day and the MDT improvement tool. Themes were identified around both positive and negative aspects of the training, summarised in Tables alongside illustrative quotes and professional roles.

Among the themes of the strength of the training, the most frequent was the ‘Quality of presentation/teaching’ (N=14), followed by ‘MDT MODE tool utility’ (N=10), and ‘Networking and collaboration’ (N=9). The evidence-based nature of the content was also widely praised, with participants valuing the integration of current research and guidelines. Several respondents noted the practical applicability of the MDT tool, particularly its utility across different professional groups. The training’s interdisciplinary relevance was emphasised, with participants reporting improved clarity in MDT roles and processes.

The two most frequently identified themes for the weakness of the training were, ‘Poor video/audio quality’ (N=18) and ‘Difficulty identifying speakers in videos’ (N=8). In addition, several respondents suggested the need **for** more practical examples and the provision of a ’toolkit’ to support implementation in clinical practice. A smaller number of participants highlighted the need for greater interactivity during sessions to enhance engagement and application of learning.

Themes identified among additional comments included ‘Desire for follow-up/implementation support’ (N=4), and ‘Request for broader content coverage’ (N=4).

## 4 Discussion

### 4.1 Summary of results

This evaluation demonstrated that structured training in multidisciplinary team (MDT) improvement, delivered by experts in implementation science and clinical practice, is well-received and valued by diverse healthcare professionals across multiple cancer specialties. Of 48 participants attending three training sessions across two cancer alliances, 48 completed the post-training survey, providing both quantitative and qualitative feedback. The quantitative analysis demonstrated high satisfaction scores, with 12 of 14 questions achieving a median response of 4/5 ("agree") to positive statements regarding the trainins, and one question achieving 4.5/5 ("agree/strongly agree"). Notably, 12 of 14 questions were answered by all participants, indicating good engagement with the evaluation itself. The qualitative analysis identified strengths in the perceived quality of presentation/teaching, utility of the assessment tool, MDT MODE tool, and opportunity for networking and collaboration among participants. However, participants also identified areas for improvement, most frequently the quality of the training videos of MDT discussions, specifically the audio quality and difficulty identifying speakers in the videos. Overall, the training successfully conveyed knowledge and skills related to MDT functioning and quality improvement, though technical delivery aspects require improvement.

### 4.2 Interpretation in light of previous evidence

The present findings align with the literature regarding the effectiveness of multidisciplinary educational interventions in healthcare. Previous research suggests that interactive, hands-on training approaches (combining didactic instruction with practical simulations) are superior to lecture-based formats in promoting knowledge retention and behavioural change[9]. The current evaluation of MDT training supports this, as participants valued the practical simulation components using the MDT MODE assessment tool and interactive teaching format. The integration of evidence-based content was also highlighted as a strength, which reflects contemporary educational best practice emphasizing the importance of grounding clinical training in rigorous research[10].

The high level of satisfaction across diverse professional groups (surgeons, radiologists, oncologists, physicians, nurses, and administrative staff) suggests that multi-disciplinary educational interventions can effectively reach and engage healthcare professionals across different career grades and specialties. This is particularly important in the cancer MDT context, where effective functioning depends on a shared understanding of the roles and contributions of all team members. The networking and collaboration theme that emerged from qualitative responses indicates that training sessions provided value beyond knowledge transfer, facilitating cross-organizational relationship-building and practice-sharing, which are important to implementing quality improvements across complex healthcare systems[11].

The modest median score of 3.5/5 ("neither agree nor disagree/agree") regarding mandatory MDT training status, however, merits further discussion. Despite high levels of satisfaction, this suggests that participants may be uncertain about making such training compulsory.

Further investigation is needed to understand this ambivalence, which may reflect concerns about implementation feasibility, resource constraints, or the need for training to be tailored to organizational context.

Although the training described above was delivered in 2019, its content has arguably become more relevant to contemporary MDT working. The courses occurred shortly prior to the publication of the NHS England guidance on streamlining MDT meetings in January 2020, which advocated that MDTs reserve full meeting discussion for complex cases, and manage straightforward cases through agreed Standards of Care, and to audit both pathways regularly. Adoption of this guidance since then has been variable. We have previously argued that successful implementation depends on teams being able to assess case complexity, the quality of the information presented and the contributions of team members, using validated tools such as MDT-MODe and the Measure of case-Discussion Complexity (MeDiC), which were the focus of the present training.[8,15–17] Moreover, the National Cancer Plan for England, published in February 2026, commits to modernising MDT working; and The Royal College of Radiologists has been tasked with a review MDT practice, with national guidance due in 2027. Delivering these reforms will require a workforce able to measure and improve team performance, equipped evidence on how training in MDT improvement is received by the professionals expected to carry it out. Our findings therefore remain directly relevant to the design of training that supports current national MDT reform. These findings include the value participants placed on practical, tool-based learning and their requests for support with implementation.

### 4.4 Strengths and limitations

This evaluation benefited from a number of methodological strengths. Firstly, the use of both quantitative and qualitative data provided complementary insights into participant experience and perceived impact. The quantitative component, with high completion rates, provided reliable descriptive data on satisfaction across multiple domains. The qualitative component employed established methods to ensure validity.

Secondly, the evaluation captured a diverse sample of healthcare professionals (N=48) across multiple cancer specialties, enhancing the generalizability of findings to broader MDT populations. The representation of surgical, medical, and allied health professionals, alongside administrative staff, reflects the genuine multidisciplinary composition of cancer MDTs and strengthens the relevance of findings to real-world MDT functioning.

Thirdly, post-training evaluation provides valuable information for course iteration and refinement. The specific identification of technical barriers (video/audio quality) and content gaps (implementation support, communication focus) offers actionable feedback for the delivery of future training courses.

Several limitations should be acknowledged. The absence of pre-training baseline measures of knowledge, attitudes, or satisfaction limits our ability to quantify the magnitude of change attributable to the training intervention. Future evaluations should incorporate pre-post designs with validated knowledge assessment instruments. Moreover, this study did not incorporate follow-up evaluation to assess sustained impact on participant knowledge, behaviour change, or subsequent MDT functioning. Our evaluation captured predominantly satisfaction, with some indication of perceived knowledge gain; however, longer-term impact is unknown. This represents an important gap, as training-to-practice transfer is not automatic and frequently requires reinforcement and organizational support.

The relatively small sample size (N=48) limits statistical power for subgroup analyses. These subgroup comparisons were also exploratory: 56 chi-squared tests were performed, and many contingency-table cells had expected counts below five. We applied a Bonferroni correction to limit the risk of false-positive findings. However, this correction is conservative, particularly when tests are correlated, as responses to related survey questions are likely to be, and it further reduces the power to detect genuine differences. Differences that did not remain significant after correction, such as the lower scores apparent for the first training day, therefore cannot be excluded, but nor should they be regarded as established. Only the differences according to grade, for knowledge of the MDT-metric and of the MDT-MODE instrument, remained significant, and even these are based on small numbers of respondents in several grade categories. All subgroup findings should therefore be regarded as hypothesis-generating. Future studies with larger sample sizes would enable more robust investigation of whether training efficacy differs across professional groups, seniority levels, or cancer specialties. Similarly, the evaluation was limited to two cancer alliances at three time points. While this provides reasonable regional representation, generalisability to other regions of the UK, or international settings is limited. Cancer MDT functioning is likely to be influenced by organizational context, resource availability, and healthcare system structure; therefore, findings should be interpreted with consideration of the UK NHS context.

### 4.4 Implications for practice, policy and research

These findings provide evidence to support the introduction of structured improvement training for cancer MDTs. The positive feedback from MDT members regarding interactive training in MDT improvement methods using the MDT MODE assessment tool suggest that this type of training might be successfully integrated into MDT development and quality improvement strategies. The identification of "Quality of presentation/teaching" as the most frequently occurring theme of strength, however, suggests that the effectiveness of training is dependent on facilitator expertise and educational approach. Cancer alliances considering this form of training should prioritise recruitment of facilitators with demonstrated expertise in both the science and practice of MDT development.

The qualitative feedback regarding need for implementation support and follow-up guidance has important practical implications. Participants requested resources and support in order to translate assessment findings into actionable improvements. This suggests that training should be seen as a component of a broader improvement support system: although necessary, as a single event it is not sufficient to lead to sustained change. Pairing training sessions with subsequent implementation support through resource provision (for example, standardised improvement templates, communication guides), regular follow-up with facilitators and the development of communities of practice may enhance the transfer of learning to practice [20].

The identification of technical delivery issues (video and audio quality) presents an opportunity for improvement of the training. Given the increasing reliance on video-based case simulations in medical education and training, investment in higher-quality multimedia production and accessibility features (e.g., subtitles, speaker identification) should be a priority in the field of MDT improvement.

Our findings may help to inform policy regarding workforce development in cancer MDT services. NHS England have tasked Cancer Alliances with facilitating MDT improvement and ensuring consistent, effective MDT functioning across regional systems [2]. The present study provides evidence that MDT members feel positively about structured, evidence-based training for MDT members, a resource that requires funding and investment in faculty training. However, the modest median response (3.5/5) regarding the suggestion that such training should be mandatory, suggests that policy frameworks should be carefully considered, perhaps with a tiered approach targeting MDT leads and designated improvement champions as first cohorts, with expansion based on demonstrated benefit[21].

This evaluation, while providing valuable insight into participant satisfaction and perceived impact, highlights important gaps in the evidence base on MDT improvement training. Future research could be augmented by including: (1) pre-post evaluation using validated outcome measures assessing knowledge and skills; (2) longer term follow-up to determine the sustainability of behavioral changes; (3) organisational outcome measurement, such as changes in MDT meeting efficiency, decision consistency, or documented patient outcomes, to establish whether individual-level training translates to team and patient-level improvement; (4) comparative effectiveness studies examining training in different formats, or for specific members of the MDT, e.g. MDT Leaders or MDT Coordinators [21,22].

## 5 Conclusion

This evaluation provides evidence supporting the value of structured, expert-led MDT improvement training delivered in an interactive, simulation-based format. High participant satisfaction across diverse professional groups, clear identification of practical value, and positive feedback regarding the educational approach suggest that further investment in this training model would be impactful. However, translating training into sustained organisational change requires additional support and investment in infrastructure and research. Future research employing robust study designs and longer-term organisational-level outcome measurement will be essential to fully establish the role of training-based interventions in improving cancer MDT functioning and ultimately patient outcomes. Additionally, optimisation of the technical delivery is needed to maximize training effectiveness.

## Data Availability

All data produced in the present work are contained in the manuscript

## Author contributions

AA: Writing – original draft, Investigation. EQ: Writing – review & editing, Methodology, Conceptualization, Investigation. BL: Investigation, Methodology, Conceptualization, Writing – review & editing. TS: Writing – review & editing, Methodology, Investigation, Writing – original draft, Conceptualization.

## Acknowledgments

The authors thank all the MDTs taking part in the training programme.

## Generative AI statement

The author(s) declared that generative AI was used in statistical analysis and editing of this manuscript.

